# Refining the Composition and Significance of Human Renal Intratubular Casts Using Spatial Protein Imaging

**DOI:** 10.1101/2025.08.20.25334083

**Authors:** Azuma Nanamatsu, Angela R. Sabo, Daria Barwinska, William S. Bowen, Jessica Hata, Michael Ferkowicz, Takashi Hato, Michael T. Eadon, Pierre C. Dagher, Avi Z. Rosenberg, Tarek M. El-Achkar, the Kidney Precision Medicine Project

## Abstract

**Background:** Renal intratubular casts are frequently observed in the distal nephron segments of the kidney and have long been regarded as a sign of renal disease. However, the composition and pathological significance of intratubular casts have remained understudied.

**Methods:** We leveraged Hematoxylin and Eosin (H&E) staining to identify intratubular casts along with concurrent Co-detection by indexing (CODEX) multiplexed spatial protein imaging on human kidney biopsy sections from the Kidney Precision Medicine Project (KPMP). We also conducted immunoblotting of Prominin-1 (PROM1) in urine and assessed its levels from publicly available urinary proteomics datasets of the KPMP consortium.

**Results:** We analyzed 424 intratubular casts across 33 individuals with kidney disease or healthy controls. We identified PROM1 and IGFBP7 as major constituents of casts (positive staining in 90.1% and 35.6%, respectively). Staining for UMOD, an established cast component, was present in 86.1%. These components exhibited distinct alterations depending on the disease state. Intratubular casts were predominantly detected in the distal nephron segments, and their presence was associated with a marked loss of NCC and AQP2 expression in the cast-containing tubular epithelium, suggesting underlying injury. The loss of these membrane transporters correlated with protein components within casts, and the presence of intra-cast PROM1 showed the strongest association, with an odds ratio of 30.8 (95% confidence interval: 13.4-71.0). Urinary PROM1 secretion was confirmed by immunoblotting and was increased in patients with acute kidney injury (AKI) compared to healthy controls (p = 0.01).

**Conclusions:** We identified PROM1, a dedifferentiation and injury marker expressed in epithelial cells, as a novel major constituent of intratubular casts. Our studies suggest that protein composition signature within casts varies with disease state and is associated with tubular injury in distal nephron segments. Our study also suggests that urinary PROM1 may serve as a biomarker for AKI.

**Key Points:**

✓ Utilized CODEX multiplex protein imaging to elucidate the intratubular cast components and the associated tubular alterations.
✓ Identified PROM1, a dedifferentiation marker, as a major constituent of intratubular casts.
✓ Protein components within casts were altered by disease state and were associated with the injury of the surrounding tubular epithelium.

## Introduction

Casts are cylindrical structures formed in the lumen of distal nephron segments of the kidney, mainly in distal convoluted tubules (DCT) and collecting ducts (CD)(1,2). Casts can comprise protein aggregates and cellular debris, including epithelial cells(1). Microscopic analysis of excreted urinary casts is a common diagnostic method in clinical nephrology. Urinary casts can be observed in healthy individuals, and an increased abundance of urinary casts is generally regarded as evidence of intrarenal pathology(3,4). Casts that are retained within kidney tubules, termed intratubular casts, may form obstructive plugs(5). While tubular obstruction with intratubular casts has been proposed to cause epithelial damage(4), the underlying composition and mechanisms are poorly defined.

Uromodulin (UMOD) is a protein synthesized in the cells of the thick ascending limb (TAL) and is the most abundant secretory protein in normal urine(6). UMOD is a major constituent of hyaline casts(2,7), the most common type of tubular casts. However, the role of UMOD in tubular cast formation and pathology remains enigmatic(8). In addition, the presence and role of other proteins in intratubular casts remain unclear. *In situ* multiplex protein expression profiling in kidney biopsy specimens is a key first step to understand intratubular cast composition and associated tubular damage.

Co-detection by indexing (CODEX) is an emerging technology for highly multiplexed spatial protein imaging using DNA-conjugated antibodies. CODEX allows simultaneous *in situ* visualization of over 60 markers, enabling protein expression profiling at a single-cell spatial resolution(9). We have established CODEX imaging and analytical pipeline on human kidney specimens(10). In this study, we utilized CODEX imaging to elucidate the spectrum of tubular cast components and the associated tubular alterations in human kidney biopsy specimens. We identified PROM1 (also known as CD133), a marker associated with injury and altered tubular cell state, as a major constituent of casts. Intratubular casts were detected mainly in distal nephron segments and were associated with a remarkable loss of NCC and AQP2 expression in the surrounding tubular epithelium, suggesting that intratubular casts contribute to the distal nephron injury.

## Methods

### Human kidney and urine specimens

Human kidney specimens collected by the KPMP consortium were acquired with informed consent and approved under a protocol by the KPMP single IRB of the University of Washington Institutional Review Board (Approval no. 20190213). Human kidney tissue biopsies were preserved in Optimal Cutting Temperature (OCT) medium. The primary adjudicated category was used to classify a subject as AKI versus CKD. KPMP manuals of procedures regarding tissue acquisition, handling recruitment sites, case adjudication and processes for tissue access are referred to in the publicly available link: https://www.kpmp.org/for-researchers. Human urine was obtained from Biopsy Biobank Cohort of Indiana under the approval of Institutional Review Board (Approval no. 1906572234)(11).

### Co-detection by indexing (CODEX/Phenocycler) multiplexed tissue imaging

CODEX imaging using its most recent Phenocycler-Fusion 2.0 (PCF) platform was performed as described previously(10), with modifications. In brief, 10-μm-thick kidney tissue sections were cut from OCT blocks onto SuperFrost Gold+ charged slides, followed by OCT removal, fixation with 1.6% PFA, and incubation with an antibody solution. Oligonucleotide probe staining and automated tile-scan imaging of the tissue between probe staining rounds were performed on the PCF fluidics handler and microscope. Images were processed as part of the PCF imaging workflow and visualized using FIJI/ImageJ. The panel used for this study includes 42 markers as recently described based on organ mapping antibody panels (10,12). Imaging data are publicly available in the KPMP atlas (www.KPMP.org).

### Hematoxylin and Eosin (H&E) staining

Flow cells were removed from post-imaging slides by soaking in xylene for four hours at room temperature. Xylene was extracted with 100% alcohol, and slides were rehydrated through sequential incubation in 95% to 70% ethanol. H&E staining was performed using Harris hematoxylin solution (HHS32, Sigma) and eosin Y solution (HT110316, Sigma).

### Cast identification and characterization

Identification of casts was conducted based on H&E staining images under the supervision of two pathologists (JH and AZR). Intratubular casts were defined as intratubular cylindrical materials, and we excluded detached cell clusters. Large casts were defined by casts with an area > 5000 µm^2^. Extrusion of cast materials into the interstitium (13) was defined by the extratubular signal of UMOD adjacent to cast-containing tubules n CODEX images. Mean fluorescence intensity of UMOD, NCC and AQP2 in tubules with or without casts was measured using freehand selections in ImageJ.

### Immunoblotting of human urine

Human urine was diluted 2x with distilled water and then denatured for 20 minutes at 60°C using 2x Sample Buffer (S3401, Sigma). Immunoblotting was conducted as described previously(14). Rabbit anti-PROM1/CD133 antibody (1:2000, 18470-1-AP, Proteintech) was used as a primary antibody. HRP-conjugated anti-IgG antibody (Proteintech) was used as a secondary antibody.

### Statistics

Statistical significance was evaluated using an unpaired t-test or Fisher’s exact test (between two conditions) and one-way ANOVA with embedded comparisons between two individual groups (among multiple conditions) with a significance level of p < 0.05.

## Results

### CODEX multiplex imaging analysis of human renal intratubular casts

To understand the protein expression profile of tubular casts and associated tubules in human kidneys, we analyzed tissue sections imaged with CODEX followed by H&E staining from 33 consecutive individuals: healthy (n=8) and with kidney disease (AKI, n=11; CKD, n=14) from the Kidney Precision Medicine Project (KPMP) consortium(15) (**Figure 1A**). Participant demographics are shown in **Figure 1B**. We used H&E staining to detect intratubular casts and identified a total of 424 intratubular casts. We identified the protein expression profile within casts, surrounding tubules and interstitium using CODEX data. Since CODEX imaging and H&E staining were conducted on the same section, we were able to perform *in situ* analysis of protein expression within casts and the surrounding tubules. The cast number, normalized per area, was trending higher in AKI and CKD patients compared to the reference samples; however, this difference did not reach statistical significance (**Figure 1C**).

**Figure 1:**
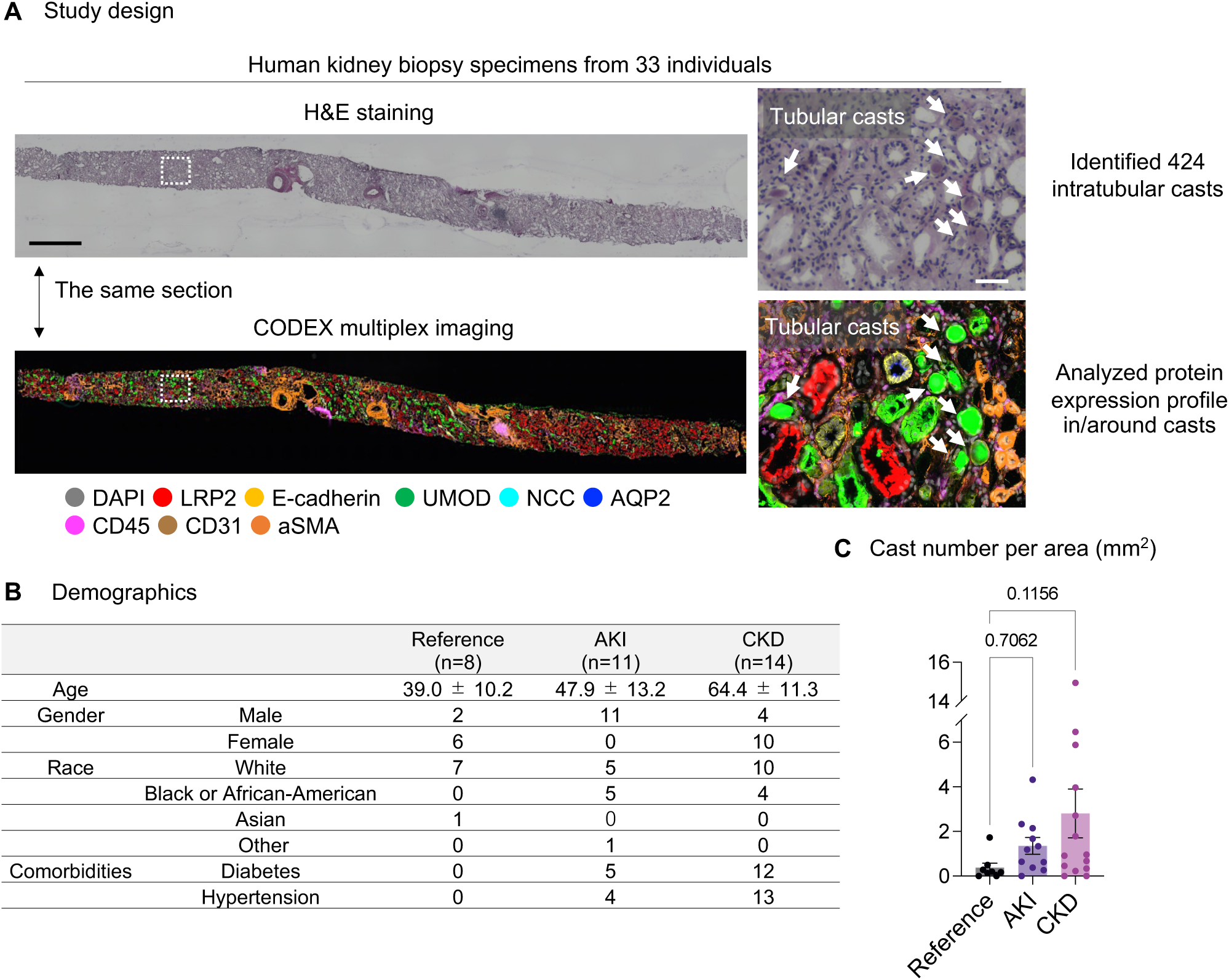
Study design. (A) We performed CODEX multiplex imaging and H&E staining on the same section of human kidney biopsy samples from the KPMP cohort. H&E staining images were used for cast identification and histological analysis. CODEX images were used to evaluate the protein expression profile of tubular casts, surrounding tubules and interstitium. Scale bars = 1mm (left panels) and 50 μm (right panels). (B) Demographics of reference and disease kidney tissue specimens. Age was presented as mean ± SD. (C) Cast number for each specimen normalized by area. Data were presented as mean ± SEM.

### UMOD, PROM1 and IGFBP7 are major components of tubular casts

We first assessed the protein expression within intratubular casts (**Figure 2A**). UMOD, an already established major component of tubular casts(2) was positive in 86.1% of casts. Notably, PROM1, a stem cell marker recently shown to be associated with dedifferentiated states in renal epithelial cells(10,15,16), was detected in 90.1% of casts. Insulin-like growth factor binding protein 7 (IGFBP7), a urinary secretory protein and a biomarker for AKI(17), was found in 35.6% of intratubular casts. Importantly, kidney injury molecule-1 (KIM-1) and vascular cell adhesion protein-1 (VCAM-1), markers associated with injury of proximal tubule (PT) cells(18,19), were rarely detected within intratubular casts (0.2% and 1.9%, respectively). These results suggest that PROM1, UMOD and IGFBP7 specifically adhere to or become incorporated into casts.

**Figure 2:**
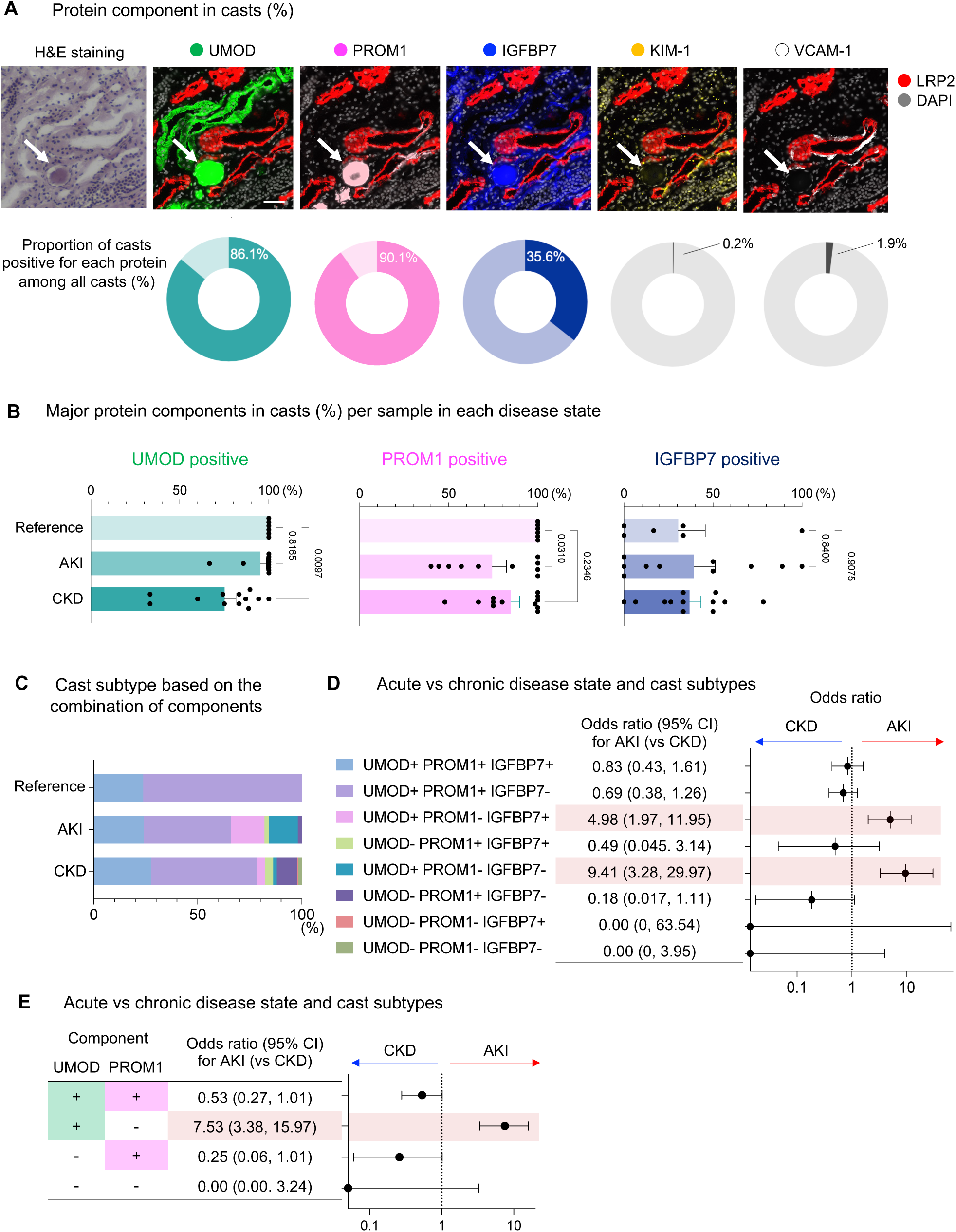
PROM1, UMOD and IGFBP7 are major components of tubular casts. (A) Representative images of casts and each protein (top). Bars = 50μm. Percentage of casts positive for each protein among all casts (bottom). (B) Ratio of PROM1-, UMOD- and IGFBP7- positive casts in each specimen in each condition. Data were presented as mean ± SEM. (C) Percent of cast subtype based on the combination of the presence or absence of PROM1, UMOD and IGFBP7 within casts in each condition. (D) Odds ratio of cast subtype (defined in (C)) for AKI versus CKD. (E) Odds ratio of cast subtype based on the presence or absence of UMOD and PROM1 for AKI versus CKD.

We next examined the positivity rate of these major components per specimen for each disease state (**Figure 2B**). PROM1 was present in all casts from reference, but its positivity rate was significantly decreased in AKI specimens. UMOD was detected in all casts in reference, but its positivity rate was reduced in CKD kidneys. The positivity of IGFBP7 in casts was comparable among conditions. These data suggest that the three major cast components behave independently in a condition-specific manner.

When casts were classified based on the presence or absence of these three major components, two major subtypes (UMOD-positive/PROM1-positive/IGFBP7-positive casts and UMOD-positive/PROM1-positive/IGFBP7-negative casts) were predominant. However, in AKI and CKD samples, additional subtypes became more prevalent, increasing the heterogeneity of cast composition (**Figure 2C**). We next reasoned that cast subtypes defined by the cast components could be associated with the stage of kidney injury (AKI or CKD). UMOD-positive/PROM-1 negative cast subtypes (UMOD-positive/PROM1-negative/IGFBP7-positive casts and UMOD-positive/PROM1-negative/IGFBP7-negative casts) were associated with higher odds of AKI rather than CKD (**Figure 2D**). We then examined whether a more simplified cast classification using UMOD and PROM1 could be used, since IGFBP7 does not appear to be a major discriminant. UMOD-positive/PROM1-negative casts were associated with higher odds of AKI rather than CKD, with an odds ratio of 7.53 (95% confidence interval: 3.38-15.97) (**Figure 2E**). The presence of PROM1-positive/UMOD-negative casts showed a nonsignificant trend toward lower odds of AKI, with an odds ratio of 0.26 (95% confidence interval: 0.06-1.01) (**Figure 2E**).

### Tubular casts are associated with loss of membrane protein expression in distal nephron segments

We next evaluated protein expression profile in cast-containing tubular epithelium to understand the localization and potential pathological role of intratubular casts. Our CODEX panel includes markers for each nephron segment (**Figure 3A**). We first assessed LRP2 (PT marker) and E-cadherin (distal nephron segment marker) expression in cast-containing tubules. Most cast-containing tubules were E-cadherin positive (**Figure 3B**), consistent with previous reports where intratubular casts were predominantly detected in the distal nephron segment (2,4). We then focused on E-cadherin-positive (distal nephron segment) tubules and assessed the detailed segmental markers: UMOD for TAL, sodium-chloride cotransporter (NCC) for DCT, and aquaporin 2 (AQP2) for CD (**Figure 3A**). In the absence of casts, 91.8% of E-cadherin-positive tubules expressed at least one of the segment-specific markers UMOD, NCC, or AQP2. Strikingly, only 10.8% of E-cadherin-positive tubules expressed any of these segment-specific markers in the presence of intratubular casts (**Figure 3C**) (p < 0.0001, Fisher’s exact test).

**Figure 3:**
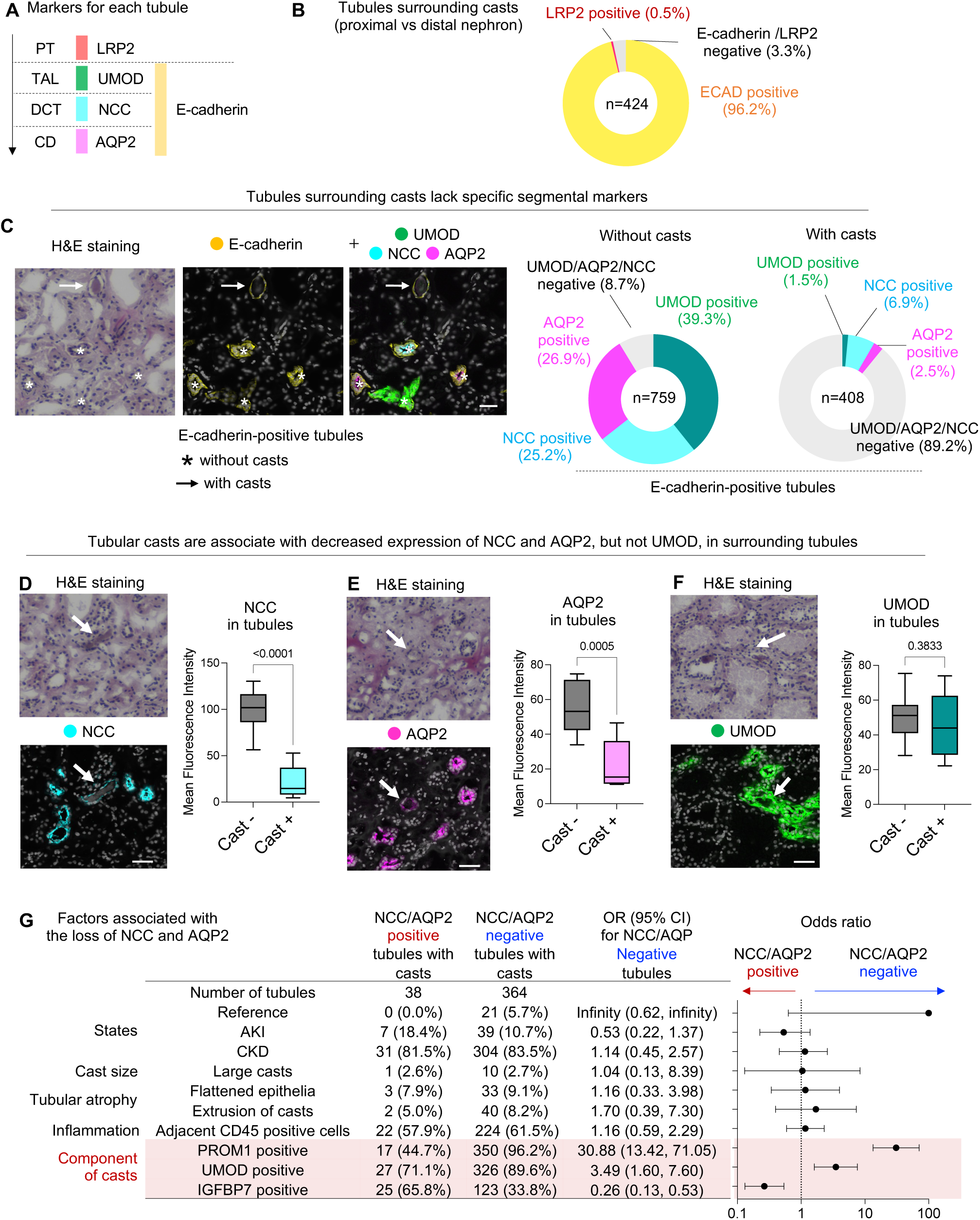
Tubular casts are associated with loss of membrane protein expression in distal nephron segments. (A) Markers for each nephron segment used in this study. PT, proximal tubule; TAL, thick ascending limb; DCT, distal convoluted tubule; CD, collecting duct. (B) Distribution of LRP2 and E-cadherin expression in tubular epithelium surrounding casts. (C) Qualitative analysis of UMOD, NCC, and AQP2 expression in E-cadherin-positive tubules with or without casts. Bars = 50μm. (D) Mean fluorescence intensity of NCC in tubules with and without casts. Bars = 50μm. n = 6-25. (E) Mean fluorescence intensity of AQP2 in tubules with and without casts. Bars = 50μm. n = 9-23. (F) Mean fluorescence intensity of UMOD in tubules with and without casts. Bars = 50μm. n = 5-14. (G) Factors associated with the loss of NCC and AQP2 expression in tubules surrounding casts.

We then focused on TAL, DCT and CD cells, which still express UMOD, NCC, or AQP2, respectively, even in the presence of intratubular casts. Expression levels of these proteins in the tubular epithelium were compared with adjacent TAL, DCT or CD epithelium without casts. We found that the NCC and AQP2 expression in DCT and CD cells were decreased in the presence of casts (**Figures 3D and E**), whereas UMOD expression in TAL epithelium was not affected by casts (**Figure 3F**). These data suggest that intratubular casts are associated with suppression of NCC and AQP2 expression in DCT and CD cells. Therefore, the UMOD/NCC/AQP2-negative tubules surrounding casts (**Figure 3C**, pie chart on the right) are likely to be explained by DCT or CD cells which have lost NCC or AQP2 expression. This is consistent with the previous findings that intratubular casts are mainly observed in the lumen of DCT and CD cells(1,2).

We next aimed to understand the factors that lead to the loss of membrane proteins in the cast-surrounding tubules in DCT and CD. To this end, we compared the characteristics of NCC/AQP2- negative and -positive tubules surrounding casts. UMOD-positive tubules were excluded from the analysis to focus on the DCT and CD epithelium. Interestingly, the odds ratio for observing NCC/AQP2-negative tubules was significantly altered by the protein components of casts, but not by disease states, cast size, flattened epithelia, extrusion of casts or adjacent inflammation (**Figure 3G**). Among the protein components, PROM1 had the strongest association with NCC/AQP2 loss in the surrounding tubules, with an odds ratio of 30.88 (95% confidence interval: 13.42-71.05).

### PROM1 is a urinary secretory protein and a potential biomarker in kidney disease

PROM1 is a transmembrane glycoprotein expressed in kidney epithelial cells, but its extracellular secretion has not been well characterized in the literature. Immunoblotting of human urine confirmed its presence in the urine (**Figure 4A**). The observed molecular weight of PROM1 was approximately 115 kDa, suggesting the excretion of full-length or near-full-length protein, rather than fragments. We assessed PROM1 expression in publicly available urinary proteomics datasets of the KPMP consortium (demographics are shown in **Figure 4B**). In this dataset, PROM1 was detected in the urine, and was significantly higher in AKI patients (p= 0.01), but not in CKD patients (p=0.12), compared to healthy reference (**Figure 4C**). Urinary KIM-1, an established biomarker for AKI(20), was also higher in AKI patients, supporting the fidelity of the proteomics analysis (**Figure 4D**).

**Figure 4:**
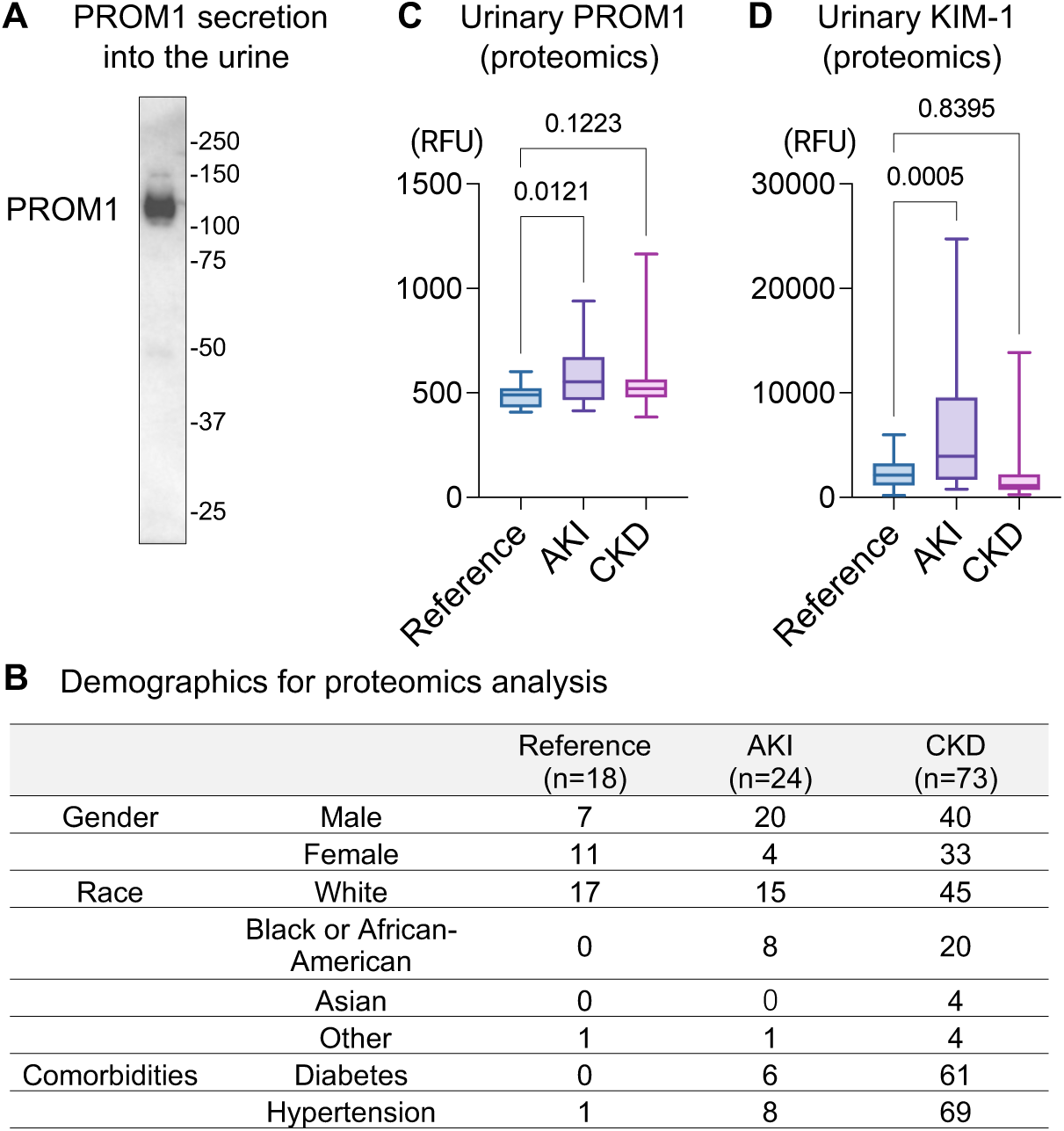
PROM1 is a urinary secretory protein and a potential biomarker in kidney disease. (A) Immunoblotting of PROM1 in human urine. (B) Demographics of reference and kidney disease individuals used for proteomics analysis. (C,D) Normalized urinary PROM1 levels (C) and KIM-1 levels (D), measured by proteomics (SomaScan), from publicly available data generated by the Kidney Precision Medicine Project. Accessed Sep 18^th^, 2024. https://www.kpmp.org. n = 18-73 per group.

## Discussion

Renal tubular casts are frequently observed in the kidneys and have long been recognized as a sign of renal disease, yet their protein composition and pathological roles have been largely unknown. In this study, we took advantage of CODEX multiplex protein imaging to investigate the intratubular cast components and associated tubular alterations in human kidneys. CODEX multiplex imaging and H&E staining were performed on the same section, enabling precise *in situ* analysis of protein expression in the casts and surrounding tubules.

We identified PROM1 as a novel major component of intratubular casts. PROM1 is a transmembrane glycoprotein expressed in kidney epithelial cells. PROM1 has been recognized as a stem cell marker(21) and has recently been linked to injury and altered cell states in renal epithelial cells(10,15,16). KIM-1 and VCAM-1 are other tubular proteins associated with injury, but they were rarely detected within casts. Therefore, the presence of PROM1 within casts is likely specific. Intra-cast PROM1 presence showed the strongest correlation with the loss of membrane proteins in the epithelium of distal nephron segments. The role of PROM1 within casts needs further investigation. We also showed that PROM1 is secreted extracellularly into the urine as a full-length or near-full-length protein. Urinary PROM1 levels were higher in AKI patients in proteomics analysis, suggesting that PROM1 may serve as a biomarker for AKI. To establish PROM1 as an AKI biomarker, it will be necessary to analyze a larger number of clinical specimens, incorporating AKI severity and time-course data, as well as the validation using complementary modalities such as ELISA. The mechanism governing the extracellular secretion of PROM1 in healthy and diseased conditions also needs to be investigated. Interestingly, PROM1-negative casts were enriched in AKI biopsy specimens, while urinary PROM1 concentration was higher in AKI patients. It is possible that AKI can alter the properties of urinary PROM1, making it less likely to be incorporated into casts and more readily excreted in the urine.

UMOD is a secretory protein in the urine and has been established as a primary constituent of hyaline casts. While UMOD plays protective roles in the urinary space against stone formation and infection(6,22), its role in intratubular/urinary casts remains unknown. We have reported that *UMOD* knockout mice still generate intratubular casts after injury, indicating that UMOD is not necessary for cast formation in mice(23). The presence of UMOD-negative intratubular casts was also reported in a mouse model with distal tubule injury (24). Here, we described the existence of UMOD-negative casts (13.9%) in human kidneys. This further supports that UMOD is not indispensable for cast formation. This study also showed that UMOD-negative casts are significantly enriched in CKD kidneys. It has been reported that all urinary casts were UMOD-positive in healthy and diseased individuals(25). Therefore, UMOD-negative casts might be more likely to be retained within the tubules rather than being excreted into the urine. This raises the hypothesis that UMOD may have a role in preventing tubular retention of casts, which could be explained by its negative charge(26) or flexibility of its polymeric form (2). A side-by-side comparison of the protein expression profiles between intratubular casts and urinary excreted casts will help to understand the protein component that may facilitate casts to pass through tubules into the urine. This study also presents the possibility that cast subtypes defined by the presence or absence of PROM1 and UMOD may serve to distinguish acute and chronic kidney disease states.

Our study provides an insight into the potential pathological role of intratubular casts. The presence of intratubular casts was associated with a substantial loss of NCC and AQP2 expression in the surrounding tubules. These data suggest that intratubular casts may contribute to injury in the distal nephron segments. We cannot exclude the possibility that loss of NCC and AQP2 precedes the tubular cast formation. That is, injured tubules could be the cause of cast retention rather than the result of it. Interestingly, protein composition within intratubular casts, but not disease state, cast size, tubular atrophy or adjacent inflammation, showed the strongest correlation with loss of membrane proteins in the distal nephron segments. How the protein composition within casts is associated with the potential distal nephron injury needs to be determined. It is possible that each protein within casts may play a role by affecting the flexibility or charge of casts.

In conclusion, this study suggests that protein composition signature within casts varies with disease state and is associated with tubular injury in distal nephron segments. Our work provides a foundation for interpreting renal pathology at the molecular level through advanced spatial analysis performed on the same histological sections.

## Limitations

First, in our workflow, histological evaluation is limited to HE staining images on fresh-frozen sections, which does not allow for further classification of intratubular casts (hyaline, granular, and waxy casts) and specific casts with established pathological significance, including light chain casts(7). Applying CODEX to formalin-fixed paraffin-embedded specimens along with H&E or PAS staining, and integrating additional histological modalities, will enable detailed histological cast classification, which can be associated with distinct protein composition within the cast. In addition, a machine learning-based method to annotate intratubular casts using Masson’s trichrome-stained images has been recently reported(1). Combining these automated methods will enable more high-throughput analysis. Second, urinary casts in the urine samples were not assessed in this study. A side-by-side comparison of the counts and protein components in urinary and intratubular casts will provide a better understanding of the molecular mechanism underlying cast plugging within tubules. Third, the expression profiles of molecules other than UMOD, NCC, and AQP2 in cells surrounding the casts remain unexplored. Spatial transcriptomic analysis will provide the gene expression landscape to define the cell state in cast-containing tubular epithelium.

## Data Availability

All data produced in the present study are available upon reasonable request to the authors

## Acknowledgments

This work was supported by National Institute of Health, National Institute for Diabetes and Digestive and Kidney Diseases R01DK111651 for TME, VA Merit Award 5I01BX003935 for TME, Dialysis Clinic Inc for TME, and Takeda Science Foundation for AN. The Kidney Precision Medicine Project (KPMP) is supported by the National Institute of Diabetes and Digestive and Kidney Diseases (NIDDK) through the following grants: U01DK133081, U01DK133091, U01DK133092, U01DK133093, U01DK133095, U01DK133097, U01DK114866, U01DK114908, U01DK133090, U01DK133113, U01DK133766, U01DK133768, U01DK114907, U01DK114920, U01DK114923, U01DK114933, U24DK114886, UH3DK114926, UH3DK114861, UH3DK114915, and UH3DK114937. We gratefully acknowledge the essential contributions of our patient participants and the support of the American public through their tax dollars. The content is solely the responsibility of the authors and does not necessarily represent the official views of the National Institutes of Health.

